# Aging metrics incorporating cognitive and physical function capture mortality risk: results from three prospective cohort studies

**DOI:** 10.1101/2021.05.14.21257213

**Authors:** Xingqi Cao, Chen Chen, Jingyun Zhang, Qian-Li Xue, Emiel O. Hoogendijk, Xiaoting Liu, Shujuan Li, Xiaofeng Wang, Yiming Zhu, Zuyun Liu

**Author notes:** These authors contributed equally to this work (Xingqi Cao, Chen Chen). Correspondence to: Xiaofeng Wang, professor, National Clinical Research Center for Aging and Medicine, Huashan Hospital, and Human Phenome Institute, Fudan University, Shanghai, China., Yiming Zhu, professor, Department of Epidemiology & Biostatistics, School of Public Health, Zhejiang University School of Medicine, 866 Yuhangtang Rd, Hangzhou, 310058, Zhejiang, China., Zuyun Liu, professor, Department of Big Data in Health Science, School of Public Health and the Second Affiliated Hospital, Zhejiang University School of Medicine, 866 Yuhangtang Rd, Hangzhou, 310058, Zhejiang, China.

## Abstract

**Background:** The aims of this study were to: 1) describe the proportions of vulnerable persons identified by three existing aging metrics that incorporate cognitive and physical function; 2) examine the associations of the three metrics with mortality; and 3) develop and validate a new simple functional score for mortality prediction.

**Methods:** The three aging metrics were the combined presence of cognitive impairment and physical frailty (CI-PF), the frailty index (FI), and the motoric cognitive risk syndrome (MCR). We operationalized them with data from two large cohort studies: the China Health and Retirement Longitudinal Study (CHARLS) and the US National Health and Nutrition Examination Survey (NHANES). Logistic regression models or Cox proportional hazard regression models, and receiver operating characteristic curves were used to examine the associations of the three metrics with mortality. A new functional score was developed and validated in the Rugao Ageing Study (RAS), an independent dataset.

**Results:** In CHARLS, the proportions of vulnerable persons identified by CI-PF, FI, and MCR were 2.2%, 16.6%, and 19.6%, respectively. Each metric predicted mortality after adjustment for age and sex, with some variations in the strength of the associations (CI-PF, odds ratio (OR)=2.87, 95% confidence interval (CI)=1.74, 4.74; FI, OR=1.94, 95% CI=1.50, 2.50; MCR, OR=1.27, 95% CI=1.00, 1.62). CI-PF and FI had additional predictive utility beyond age and sex, as demonstrated by integrated discrimination improvement, and continuous net reclassification improvement (all P <0.001). These results were replicated in NHANES. Furthermore, we developed a new functional score by selecting six self-reported items from CI-PF and FI in CHARLS, and demonstrated that it predicted mortality risk. This functional score was further validated in RAS. To facilitate the quick screening of persons with deteriorations in cognitive and physical function, we introduced a publicly available online tool designed for this new functional score.

**Conclusions:** Despite the inherent differences in the aging metrics incorporating cognitive and physical function, they consistently capture mortality risk. The findings support the incorporation of cognitive and physical function for risk stratification in both Chinese and US persons, but call for caution when applying them in specific study settings.

## Introduction

The aging process is characterized by deteriorating function across a broad spectrum of physiological systems over time. To quantify the complex aging process, aging metrics have been developed in molecular, phenotypic, and functional domains [1]. Functional metrics of aging include cognitive and physical function. It has been observed that age-related decline in cognitive and physical function coexists in many older persons [2], implying possible shared mechanisms underlying the two functional aspects [2]. Furthermore, older persons have increased risk of poor prognosis (e.g., disability [3, 4], death [4-8]) when having problems in cognitive and physical function simultaneously. The potential link between cognitive and physical function motivated many researchers to explore aging metrics that incorporate the two functional aspects [9-11]. Such composite aging metrics could serve as a new target for preventing or delaying the onset of the disability and extending life expectancy in older persons [12, 13].

To date, there are three main aging metrics reported in the literature (**Figure 1A**, and **Table S1**, see for details [14]). First, in 2013, an International Association of Gerontology and Geriatrics (IAGG) consensus group proposed cognitive frailty as the simultaneous presence of both cognitive impairment and physical frailty (PF) in non-demented older persons [15] (referred to as CI-PF). PF represents a state of increased vulnerability to stressor resulting from the cumulative decline in multiple physiological systems [16, 17]. Second, the frailty index (FI) integrates deficits across multiple domains including cognitive and physical function, which results in a score reflecting risks across various outcomes (e.g., hospitalization, death) [18]. Finally, Verghese et al [19] proposed the motoric cognitive risk syndrome (MCR), characterized by the simultaneous presence of subjective cognitive complaints and slow gait. The latter has been widely used to operationalize physical function [20]. Despite some conceptual overlap in the three metrics above, there are substantial differences in their operationalizations and characteristics. The incomplete understanding (e.g., predictiveness) of these metrics has hampered their utility in research and clinical practice.

**Figure 1.**
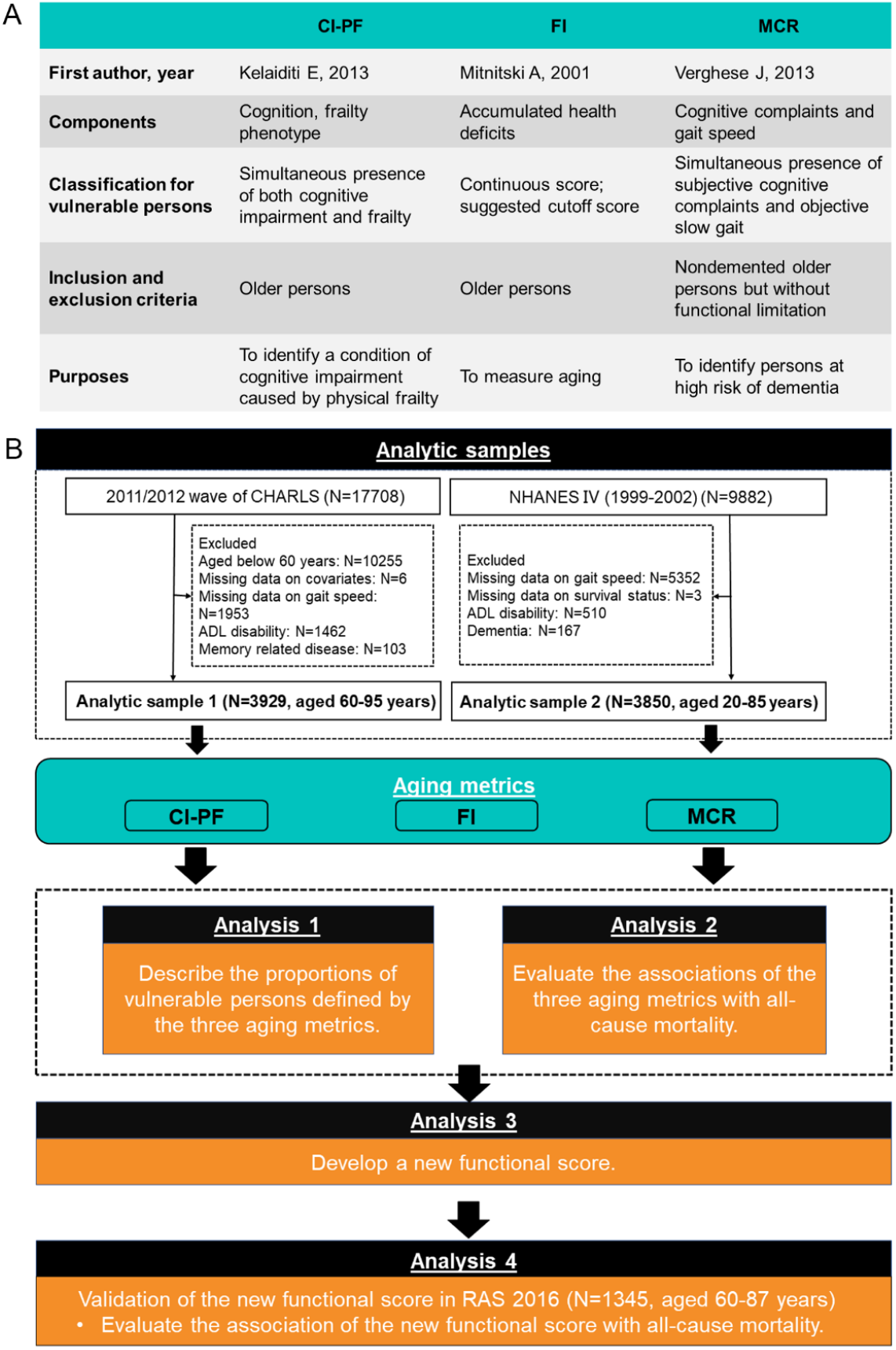
Roadmap for the comprehensive analyses of the three aging metrics incorporating cognitive and physical function. Notes: CHARLS, China Health and Retirement Longitudinal Study; NHANES, National Health and Nutrition Examination Survey; CI-PF, cognitive impairment and physical frailty; FI, frailty index; MCR, Motoric Cognitive Risk syndrome. A describes the details on the three aging metrics incorporating cognitive and physical function. B shows the assembly of analytic samples and the detailed analyses performed in this study.

In this study, we performed comprehensive analyses to describe the three metrics using data from two national prospective cohort studies: the China Health and Retirement Longitudinal Study (CHARLS) and the US National Health and Nutrition Examination Survey (NHANES). We first described the proportions of persons identified as vulnerable by the three aging metrics that incorporate cognitive and physical function. Next, we examined the associations of the three metrics with mortality. Building on the observations (i.e., the relatively better performance of CI-PF and FI than MCR in mortality prediction), we then developed a new simple functional score that integrated CI-PF and FI. Finally, we validated the new functional score in another prospective cohort study from China, Rugao Ageing Study (RAS, an independent dataset). To further facilitate the quick screening of vulnerable persons, we introduced a publicly available online tool designed for this new functional score.

## Methods

### Study population

Persons in CHARLS were first recruited in 2011/2012, and completed three follow-up visits biennially up to 2017/2018. The CHARLS was approved by the Biomedical Ethics Review Committee of Peking University, and all persons provided informed consent. As shown in **Figure 1B**, out of 17708 persons aged 45 years and older enrolled in the baseline survey (2011/2012), we excluded those aged below 60 years (N=10255; because gait speed was measured only in persons aged 60 years and over), with missing data on covariates (N=6), missing data on gait speed (N=1953), who had disability in activities of daily living (ADL) (N=1462), or had the memory-related disease (e.g., Alzheimer’s disease, brain atrophy, and Parkinson’s disease) (N=103), leaving the analytic sample 1 of 3929 persons aged 60-95 years. Persons in NHANES were first recruited in the 1999-2002 cycle. The NHANES was approved by the National Center for Health Statistics Research Ethics Review Board, and all persons provided informed consent. Out of the 9882 persons aged 20 years and older, we excluded persons with missing data on gait speed (N=5352; gait speed measurement is needed to construct both PF and MCR, but from 2003-2004 cycle on, gait speed was no longer captured in NHANES) or final mortality status (N=3), who had disability in ADL (N=510), or had dementia (N=167), leaving the analytic sample 2 of 3850 persons aged 20-85 years. In RAS, 1345 persons aged 60-87 years from the 2016 wave were included for validation of the new simple functional score in this study. The RAS was approved by the Human Ethics Committee of the School of Life Science at Fudan University, and all persons provided informed consent. A more detailed description of the study population is provided in **Supplementary Materials**.

### Measures

#### Cognitive function

In CHARLS, cognitive function was assessed by three tests, including the Telephone Interview of Cognitive Status-10 (TICS-10), word recall, and figure drawing, with higher scores indicating better cognitive function (range: 0-21) [21]. According to the literature [22], a person was classified as having cognitive impairment if the summary score fell more than 1 standard deviation (SD) below age-appropriate norms.

During the 1999-2002 cycle, NHANES used the Digit Symbol Substitution Test (DSST) to assess cognitive function, with higher scores indicating better cognitive function (range: 0-133). According to the literature [23], a person was classified as having cognitive impairment if the summary score was below the median DSST score (i.e., 40).

Cognitive function in RAS was assessed using the Hasegawa Dementia Scale-Revised (HDS-R). The HDS-R measures five domains of cognitive function including orientation, memory function, common sense, calculation, and immediate memory, with higher scores indicating better performance (range: 0-32.5) [24]. According to the literature [25], a person was classified as having cognitive impairment if the summary score was 21.5 or below.

#### Aging metrics incorporating cognitive and physical function

As mentioned above (and in **Figure 1A**), we considered three metrics incorporating cognitive and physical function: CI-PF, FI, and MCR.

#### CI-PF

The CI-PF was defined as the simultaneous presence of both cognitive impairment and PF in non-demented older persons, proposed in 2013 by an international consensus group) [15]. PF was measured using the Fried frailty phenotype approach [26], and had been previously developed and validated in the CHARLS [27] and NHANES [28], respectively. Persons were classified as frail if they met ≥3 of the five items; otherwise, they were classified as non-frail. Based on the two components, i.e., cognitive impairment and PF, we defined four combined groups as done in previous studies [11]: normal cognition & non-frailty, cognitive impairment & non-frailty, normal cognition & frailty, and cognitive impairment & frailty. The cognitive impairment and frailty group was defined as vulnerable.

#### FI

The FI was based on the degree of accumulation of health deficits and represented an alternative instrument of frailty that incorporates many health dimensions (e.g., comorbidities and disabilities) including cognition [18]. The FI score was calculated as a ratio of the number of deficits in a person out of the total possible deficits considered [29], with a range of 0 to 1 (see details in **Supplementary Material**). We categorized FI into three groups, based on the widely used cut-off values [30]. A FI ≤0.10 was considered as non-frail, 0.10< FI ≤0.21 was pre-frail, and FI >0.21 was frail. The group with frailty was defined as vulnerable.

#### MCR

The MCR was defined as the simultaneous presence of both subjective cognitive complaints and objective slow gait, in the absence of a diagnosis of dementia and disability in ADL [31]. The group of persons with MCR was defined as vulnerable.

#### Mortality

The death information in CHARLS was collected from the exit interview in 2013, 2015, and 2018 waves. Because the exact date of death was not available in the 2015 and 2018 waves, we constructed a binary variable to denote the occurrence of death within the 6-year follow-up since baseline in this study. All-cause mortality during approximately 13.8-year follow-up in NHANES was based on linked data from records taken from the National Death Index through December 31, 2015, provided through the Centers for Disease Control and Prevention. Death information in RAS was collected from the Funeral home of Rugao and Rugao Civil Affairs Bureau. The village or community doctors were responsible to investigate and validate the cause of death.

#### Covariates

We considered covariates including age, sex, education, and chronic disease, as detailed in **Supplementary Material**.

### Statistical analyses

The analytic plan for this study was described in **Figure 1B**. In analysis 1, we first described the characteristics of the full sample of CHARLS and NHANES, as well as the characteristics of vulnerable persons defined by the three metrics using mean (±SD) or counts (percentages). We then plotted the proportions of vulnerable persons identified by the three aging metrics at baseline in the full sample, as well as stratified by age categories (<65 years, and ≥65 years) and sex. To check the consistency of the three metrics, we further presented the distribution of persons with CI-PF and MCR across the FI groups.

In analysis 2, we evaluated the associations of the three metrics with all-cause mortality. We used logistic regression models in CHARLS (because the exact timing of death during the follow-up period was unknown in CHARLS) and Cox proportional hazard regression models in NHANES. Model 1 adjusted for age and sex. Model 2 additionally adjusted for education, and residence (CHARLS) or ethnicity/race (NHANES). For logistic regression models, we documented odds ratios (ORs) and corresponding 95% confidence intervals (CIs). For Cox proportional hazard regression model, we documented hazard ratios (HRs) and corresponding 95% CIs. Then, receiver operating characteristic (ROC) curves were used to evaluate the utility of the three metrics for mortality prediction beyond basic models with age and sex, in both CHARLS and NHANES. Indices including the delta C-statistic, integrated discrimination improvement (IDI), and continuous net reclassification improvement (NRI) were calculated, in comparison to that of the basic model. Delta C-statistic equals to x% means that the difference in predicted risks between the persons with and without the outcome increased by x% in the updated model. IDI equals to x% means that the difference in average predicted risks between the persons with and without the outcome increased by x% in the updated model. Continuous NRI equals to x% means that compared with persons without outcome, persons with outcome were almost x% more likely to move up a category than down.

Building on the findings that CI-PF and FI performed relatively better than MCR, we then constructed a new simple functional score that integrated CI-PF and FI (using items from the two metrics) following the procedures previously described [32] (analysis 3, details in **Supplementary Materials**). Considering that several self-reported diseases (e.g., chronic lung disease, heart disease) items were retained in the stepwise logistic regression models, we replaced them with one disease count variable. After carefully screening self-reported items for mortality prediction and their properties (e.g., reflect cognitive or physical function), we included one item for cognition (i.e., serial subtraction of 7 from 100) and five items of physical function (i.e., having a body mass index (BMI) of 18.5 kg/m^2^ or less, disease count, and having limitations in running/jogging 1 km, walking 1 km and climbing several flights of stairs) to develop the new functional score. ROC curves were then used to evaluate the association of the new functional score with mortality risk in CHARLS. We calculated the delta C-statistic, IDI, and continuous NRI in comparison to that of the basic model with age and sex. Finally, we validated the new functional score for all-cause mortality prediction in RAS (analysis 4). ROC curves, the delta C-statistic, IDI, and continuous NRI were used, in comparison to that of the basic model with age and sex.

All statistical analyses were performed using R version 3.6.3 (2020-02-29) and SAS version 9.4 (SAS Institute, Cary, NC). A P value of <0.05 (two-tailed) was considered statistically significant.

## Results

### The characteristics of the study population

As shown in **Table 1**, the mean ages of the 3929 persons in CHARLS and the 3850 persons in NHANES were 67.4 (SD=6.3) years and 65.6 (SD=9.6) years, respectively. The proportions of males were 53.5% in CHARLS and 50.1% in NHANES. In RAS, the mean age of the 1345 persons was 77.2 (SD=3.9) years, and the proportion of males was 46.4% (**Table S2**).

**Table 1.** Summary characteristics of the total sample and for vulnerable persons identified by the three aging metrics, CHARLS 2011/2012 and NHANES 1999–2002.

| Characteristics | CHARLS |  |  |  | NHANES |  |  |  |
| --- | --- | --- | --- | --- | --- | --- | --- | --- |
|  | Total | Vulnerable persons |  |  | Total | Vulnerable persons |  |  |
|  |  | CI-PF | FI | MCR |  | CI-PF | FI | MCR |
| N | 3929 | 85 | 653 | 770 | 3850 | 104 | 1035 | 167 |
| Age, mean±SD | 67.4±6.3 | 73.2±7.8 | 68.7±7.1 | 66.0±5.4 | 65.6±9.6 | 72.0±7.3 | 69.6±9.3 | 71.6±9.4 |
| Male, % | 2102 (53.5) | 28 (32.9) | 269 (41.2) | 319 (41.4) | 1927 (50.1) | 46 (44.2) | 468 (45.2) | 83 (49.7) |
| Residence, rural, % | 2427 (61.8) | 65 (76.5) | 435 (66.6) | 533 (69.2) | — | — | — | — |
| Race/Ethnicity <sup>a</sup>, % |  |  |  |  |  |  |  |  |
| Non-Hispanic white | — | — | — | — | 2130 (56.6) | 47 (46.1) | 545 (53.6) | 84 (51.5) |
| Non-Hispanic black | — | — | — | — | 673 (17.9) | 20 (19.6) | 194 (19.1) | 23 (14.1) |
| Hispanic | — | — | — | — | 957 (25.5) | 35 (34.3) | 278 (27.3) | 56 (34.4) |
| Education <sup>b</sup>, % |  |  |  |  |  |  |  |  |
| Category 1 | 1296 (33.0) | 61 (71.8) | 269 (41.2) | 334 (43.4) | 1496 (39.0) | 71 (68.3) | 526 (51.0) | 90 (54.9) |
| Category 2 | 1859 (47.3) | 24 (28.2) | 297 (45.5) | 352 (45.7) | 868 (22.6) | 17 (16.3) | 213 (20.7) | 23 (14.0) |
| Category 3 | 511 (13.0) | 0 | 60 (9.2) | 71 (9.2) | 799 (20.8) | 8 (7.7) | 184 (17.8) | 28 (17.1) |
| Category 4 | 195 (5.0) | 0 | 19 (2.9) | 11 (1.4) | 677 (17.6) | 8 (7.7) | 108 (10.5) | 23 (14.0) |
| Category 5 | 68 (1.7) | 0 | 8 (1.2) | 2 (0.3) | — | — | — | — |
| Disease counts <sup>c</sup>, % |  |  |  |  |  |  |  |  |
| 0 | 1116 (28.4) | 23 (27.1) | 40 (6.0) | 173 (22.5) | 805 (20.9) | 1 (1.0) | 14 (1.4) | 12 (7.2) |
| 1 | 1252 (31.9) | 30 (35.3) | 107 (16.0) | 252 (32.7) | 1138 (29.6) | 17 (16.3) | 126 (12.2) | 20 (12.0) |
| 2 | 885 (22.5) | 17 (20.0) | 166 (24.8) | 178 (23.1) | 961 (25.0) | 19 (18.3) | 244 (23.6) | 47 (28.1) |
| 3 | 426 (10.8) | 12 (14.1) | 169 (25.3) | 100 (13.0) | 574 (14.9) | 28 (26.9) | 327 (31.6) | 37 (22.2) |
| ≥4 | 250 (6.4) | 3 (3.5) | 187 (28.0) | 67 (8.7) | 372 (9.7) | 39 (37.5) | 324 (31.3) | 51 (30.5) |
| Cognitive impairment, % | 1348 (34.3) | / | 307 (47.0) | 397 (51.6) | 1055 (27.4) | / | 411 (39.7) | 70 (41.9) |
| Physical frailty, % | 165 (4.2) | / | 78 (11.9) | 78 (10.1) | 338 (8.8) | / | 302 (29.2) | 73 (43.7) |
| Gait speed, mean±SD | 0.7±0.3 | 0.4±0.3 | 0.6±0.3 | 0.5±0.1 | 6.9±3.0 | 11.4±5.0 | 8.7±4.1 | 8.8±5.0 |
| Slow gait, % | 1979 (50.4) | 70 (82.4) | 396 (60.6) | / | 1252 (32.5) | 71 (68.3) | 449 (43.4) | / |
| Cognitive complaints, % | 1428 (36.4) | 54 (63.5) | 381 (58.4) | / | 409 (10.6) | 28 (26.9) | 289 (27.9) | / |
| The cognition score<sup>d</sup>,<br>mean±SD | 9.8±4.3 | 3.9±2.2 | 8.3±4.3 | 8.0±4.1 | 42.0±14.7 | 23.2±10.7 | 37.9±14.3 | 35.5±12.8 |

Notes: CHARLS, China Health and Retirement Longitudinal Study; NHANES, National Health and Nutrition Examination Survey; CI-PF, cognitive impairment and physical frailty; FI, frailty index; MCR, Motoric Cognitive Risk syndrome; SD, standard deviation. CI-PF, FI, and MCR indicate vulnerable persons identified by the three aging metrics, as described in Methods.
<sup>a</sup> Ninety persons who self-identified as other races (including multi-racial) were excluded.
<sup>b</sup> Ten persons with missing data on education were excluded. In CHARLS, category 1 to 5 indicates “illiteracy”, “elementary”, “middle”, “senior” and “college and higher than college”, respectively; In NHANES, category 1 to 4 indicates “lower than high school”, “high school or general educational development”, “some college”, and “college”, respectively.
<sup>c</sup> In CHARLS, chronic diseases included hypertension, diabetes or high blood sugar, cancer or malignant tumor, chronic lung disease, heart problems, stroke, kidney disease, stomach or other digestive diseases, arthritis or rheumatism, and asthma. In NHANES, chronic diseases included congestive heart failure, stroke, cancer, chronic bronchitis, emphysema, cataracts, arthritis, type 2 diabetes, hypertension, and myocardial infarction.
<sup>d</sup> In CHARLS, the cognitive function was assessed by three tests, including the Telephone Interview of Cognitive Status-10 (TICS-10), word recall, and figure drawing. In NHANES, the cognitive function was assessed by the Digit Symbol Substitution Test.

### How many persons are identified as vulnerable by the three metrics combining cognitive and physical function?

As shown in **Figure 2**, we observed large variations in the proportions of vulnerable persons using the three metrics in CHARLS and NHANES. In CHARLS, the proportions of vulnerable persons identified by CI-PF, FI, and MCR were 2.2%, 16.6%, and 19.6%, respectively (**Table S3** and **Figure 2**). In NHANES, the proportions of vulnerable persons identified by CI-PF, FI, and MCR were 2.7%, 26.9%, and 4.3%, respectively. We further presented the distribution of persons with CI-PF and MCR across the FI groups in CHARLS and NHANES in **Table S3**. We found that in CHARLS, persons who were non-frail and pre-frail defined by FI, 26.2% and 36.2% belonged to the cognitive impairment & non-frailty group for CI-PF, and 10.6% and 23.3% were classified as MCR.

**Figure 2.**
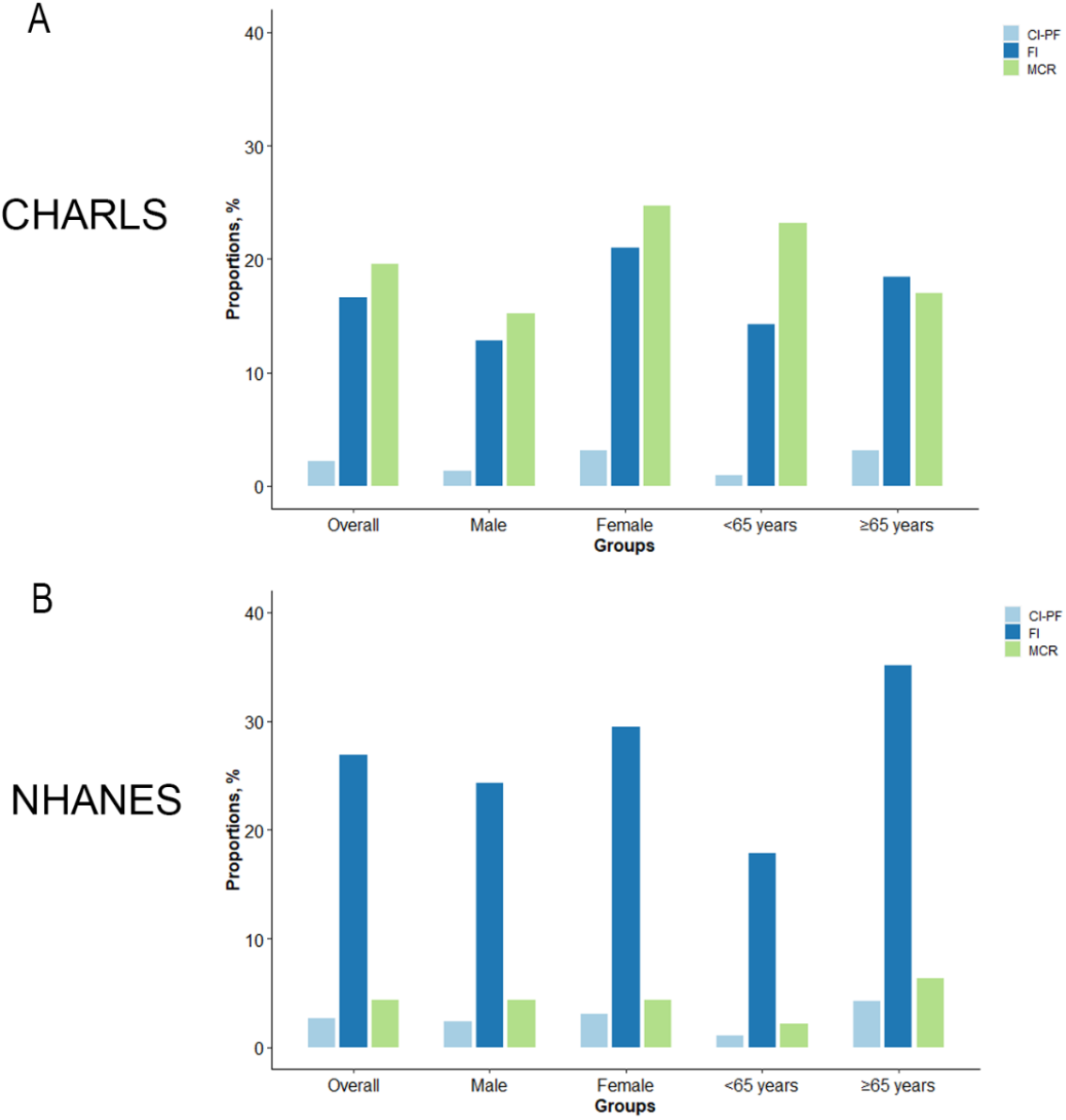
Proportions of vulnerable persons identified by the three aging metrics incorporating cognitive and physical function in CHARLS (A) and NHANES (B). Notes: CHARLS, China Health and Retirement Longitudinal Study; NHANES, National Health and Nutrition Examination Survey; CI-PF, cognitive impairment and physical frailty; FI, frailty index; MCR, Motoric Cognitive Risk syndrome.

There were also differences in characteristics among the three groups defined as vulnerable. For example, in CHARLS, the mean age of persons was 73.2 (SD=7.8) years for those defined as vulnerable according to CI-PF, for the FI this was 68.7 (SD=7.1) years, and for MCR this was 66.0 (SD=5.4) years. In all vulnerable groups, proportions of males (32.9% for CI-PF, 41.2% for FI, and 41.4% for MCR) were lower than that of females (**Table 1**).

### Do aging metrics incorporating cognitive and physical function predict mortality?

**Table 2** presents the associations of the three metrics with all-cause mortality. Overall, we found consistent and significant associations regardless of study cohorts (CHARLS and NHANES) and metrics (CI-PF, FI, and MCR), but there were some variations in the strength of the associations. When using the CI-PF, compared with the normal cognition & non-frailty group, the multivariable-adjusted ORs or HRs of the cognitive impairment & non-frailty group, normal cognition & frailty group, and cognitive impairment & frailty group for all-cause mortality were 1.35 (95% CI=1.08, 1.69), 1.69 (95% CI=0.99, 2.90), and 2.42 (95% CI=1.46, 4.02) in CHARLS (as previously reported [11]), and 1.39 (95% CI=1.23, 1.57), 3.09 (95% CI=2.58, 3.69), and 2.78 (95% CI=2.19, 3.54) in NHANES, respectively. When using the FI, compared with the non-frail group, the multivariable-adjusted ORs or HRs of the pre-frailt group and the frail group were 1.27 (95% CI=1.02, 1.57), and 1.82 (95% CI=1.41, 2.35) in CHARLS, and 1.36 (95% CI=1.18, 1.57), and 2.58 (95% CI=2.23, 2.98) in NHANES, respectively. When using the MCR, compared with persons without MCR, the multivariable-adjusted OR or HR of persons with MCR were 1.16 (95% CI=0.91, 1.47) in CHARLS, and 1.83 (95% CI=1.51, 2.22) in NHANES, respectively. Moreover, we found that vulnerable persons, as defined by the three aging metrics, had a much steeper decline in survival over approximately 13.8 years of follow-up in NHANES (**Figure S1**).

**Table 2.** Associations of the three aging metrics incorporating cognitive and physical function with all-cause mortality.

| Aging metrics |  | Model 1 | Model 2 |
| --- | --- | --- | --- |
| CHARLS |  | OR (95% CI) | OR (95% CI) |
| CI-PF | Normal cognition & non-frailty | Ref | Ref |
|  | Cognitive impairment & non-frailty | 1.59 (1.29, 1.96) | 1.35 (1.08, 1.69) |
|  | Normal cognition & frailty | 1.88 (1.10, 3.20) | 1.69 (0.99, 2.90) |
|  | Cognitive impairment & frailty | 2.87 (1.74, 4.74) | 2.42 (1.46, 4.02) |
| FI | Non-frail | Ref | Ref |
|  | Pre-frail | 1.34 (1.08, 1.66) | 1.27 (1.02, 1.57) |
|  | Frail | 1.94 (1.50, 2.50) | 1.82 (1.41, 2.35) |
| MCR | Absence | Ref | Ref |
|  | Presence | 1.27 (1.00, 1.62) | 1.16 (0.91, 1.47) |
| NHANES |  | HR (95% CI) | HR (95% CI) |
| CI-PF | Normal cognition & non-frailty | Ref | Ref |
|  | Cognitive impairment & non-frailty | 1.39 (1.25, 1.56) | 1.39 (1.23, 1.57) |
|  | Normal cognition & frailty | 3.16 (2.66, 3.76) | 3.09 (2.58, 3.69) |
|  | Cognitive impairment & frailty | 2.85 (2.26, 3.60) | 2.78 (2.19, 3.54) |
| FI | Non-frail | Ref | Ref |
|  | Pre-frail | 1.39 (1.21, 1.60) | 1.36 (1.18, 1.57) |
|  | Frail | 2.68 (2.33, 3.10) | 2.58 (2.23, 2.98) |
| MCR | Absence | Ref | Ref |
|  | Presence | 1.91 (1.58, 2.31) | 1.83 (1.51, 2.22) |

Notes: CHARLS, China Health and Retirement Longitudinal Study; OR, odds ratio; CI, confidence interval; CI-PF, cognitive impairment and physical frailty; FI, frailty index; MCR, Motoric Cognitive Risk syndrome; NHANES, National Health and Nutrition Examination Survey; HR, hazard ratio. As previously reported (Chen et al., 2020), significant differences in all-cause mortality between the four CI-PF groups were observed. As described in the Methods section, the date of death was not provided in the 2015 and 2017 waves of CHARLS. Thus, we included a binary variable to denote the occurrence of death over the 6-year follow-up since baseline in this study. We then used logistic regression models to examine the associations of the three aging metrics with death and documented ORs (95% CIs) in CHARLS. Since the date of death was available in NHANES, we used Cox proportional hazard regression methods and documented HRs (95% CIs) in NHANES. Model 1: adjusted for age, and sex; Model 2: adjusted for age, sex, education, and residence (in CHARLS) or ethnicity/race (in NHANES).

As shown in **Figure 3**, we examined the ROC curves using various models, such as the basic model with age and sex only, and the basic model with or without the three metrics included. We found that in almost all cases, the three metrics added predictive utility, except for MCR in CHARLS. The results suggested that aging metrics incorporating cognitive and physical function capture something above and beyond what can be explained by age and sex when predicting mortality. Compared with the basic model (with age and sex only), the models including CI-PF or FI (only in NHANES) had better discrimination ability, as demonstrated by significant increases in C-statistics that ranged from 0.006 to 0.032 (**Table 3**). The better performance of CI-PF and FI was further demonstrated by significant improvements in reclassification as assessed by IDI (range: 0.009-0.043) and continuous NRI (range: 0.155-0.568). Additionally, we performed sensitivity analyses in which we excluded persons aged below 60 years (N=1099) in NHANES and found that the results remain unchanged (**Table S4** and **Figure S2**).

**Table 3.** The reclassification performance and improvement in discrimination of aging metrics incorporating cognitive and physical function.

| Aging metrics | Delta C-statistic | P value | IDI | P value | NRI | P value |
| --- | --- | --- | --- | --- | --- | --- |
| **CHARLS** |  |  |  |  |  |  |
| CI-PF | 0.008 | 0.017 | 0.011 | <0.0001 | 0.211 | <0.001 |
| FI | 0.006 | 0.130 | 0.009 | <0.0001 | 0.155 | <0.001 |
| MCR | 0.0003 | 0.843 | 0.001 | 0.035 | -0.027 | 0.441 |
| **NHANES** |  |  |  |  |  |  |
| CI-PF | 0.021 | <0.001 | 0.033 | <0.001 | 0.568 | <0.001 |
| FI | 0.029 | <0.001 | 0.043 | <0.001 | 0.332 | <0.001 |
| MCR | 0.003 | 0.033 | 0.007 | <0.001 | 0.125 | <0.001 |

Notes: IDI, integrated discrimination improvement; NRI, net reclassification index; CHARLS, China Health and Retirement Longitudinal Study; CI-PF, cognitive impairment and physical frailty; FI, frailty index; MCR, Motoric Cognitive Risk syndrome; NHANES, National Health and Nutrition Examination Survey. We calculated the IDI and continuous NRI using the R package “PredictABEL”, in comparison to that of the model with age and sex. NRIs equal to x% means that compared with persons without outcome, persons with outcome were almost x% more likely to move up a category than down. IDI equals to x% means that the difference in average predicted risks between the persons with and without the outcome increased by x% in the updated model.

**Figure 3.**
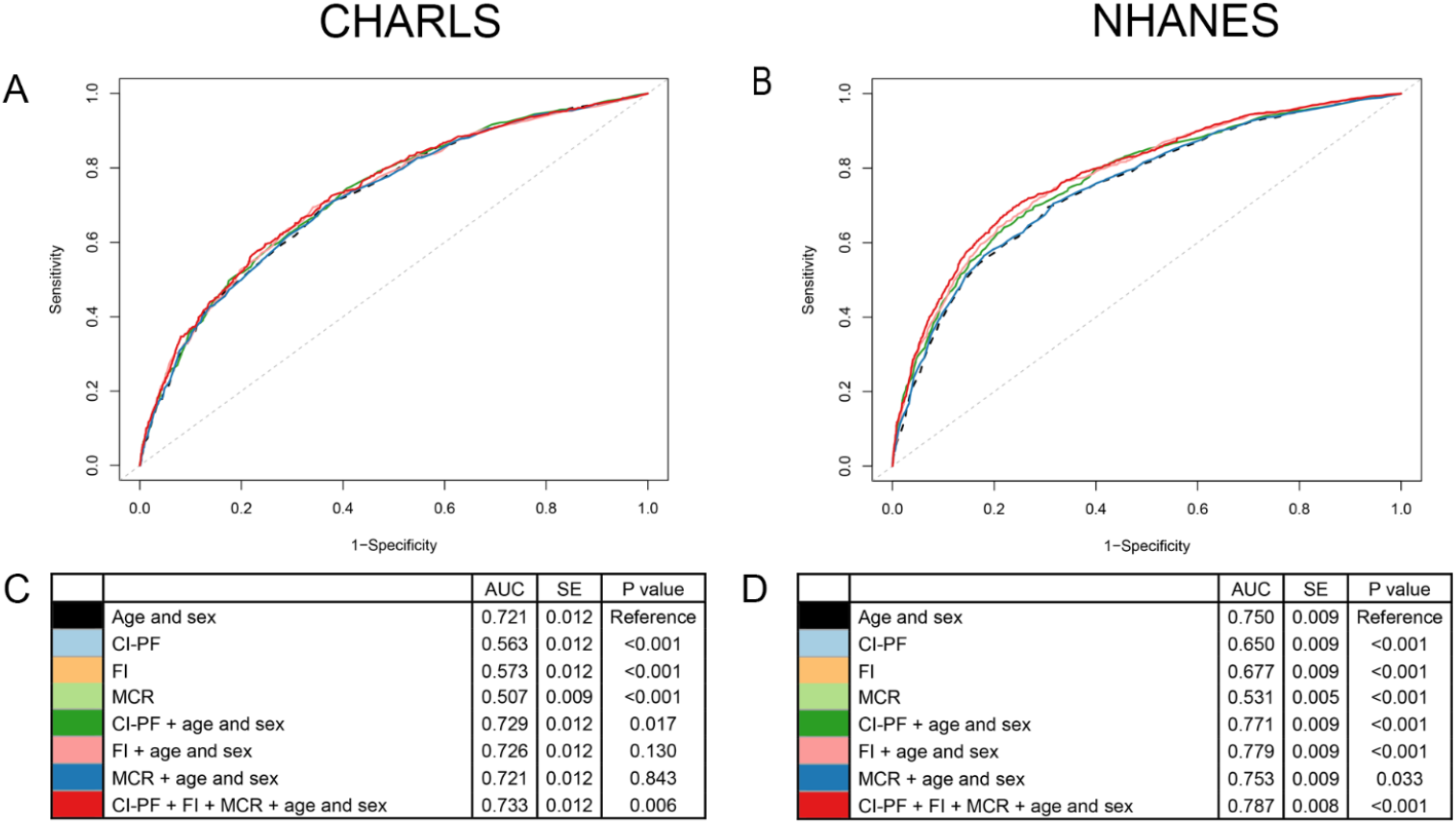
Receiver-operator characteristics (ROC) curves for prediction of all-cause mortality in CHARLS and NHANES. Notes: CHARLS, China Health and Retirement Longitudinal Study; NHANES, National Health and Nutrition Examination Survey; CI-PF, cognitive impairment and physical frailty; FI, frailty index; MCR, Motoric Cognitive Risk syndrome; AUC, area under the curve; SE, standard error. A and B show ROC curves for the prediction of all-cause mortality. C and D show the AUC for each model. A and C are based on the CHARLS. B and D are based on the NHANES.

### Can we develop a new simple functional score based on self-reported items to predict mortality?

Building on the observations above (i.e., the relatively better performance of CI-PF and FI than MCR in mortality prediction), we developed a new simple functional score that integrated CI-PF and FI in CHARLS. The components of the new functional score in CHARLS were presented in **Table 4**, and the total score ranged from 0 to 20. The estimation of death risk for the functional score was presented in **Table S5**. The area under the curve (AUC) of the model with the functional score, age, and sex was 0.742 (**Figure 4B**). The results suggested that the new functional score can predict mortality risk. The components of the new functional score in NHANES were presented in **Table S6**, and the total score ranged from 0 to 28.

**Table 4.** Components of the new functional score in CHARLS.

| Components | Construction |  | Person #1 |  |
| --- | --- | --- | --- | --- |
|  | Category | Risk points | Response | Points |
| Serial subtraction of 7 from 100 | 0 | 4 |  |  |
|  | 1 | 3 |  |  |
|  | [2, 3] | 2 | 2 | 2 |
|  | 4 | 1 |  |  |
|  | 5 | 0 |  |  |
| Having a body mass index (BMI) of 18.5 kg/m<sup>2</sup> or less | No | 0 | No | 0 |
|  | Yes | 5 |  |  |
| Disease count | [0, 1] | 0 |  |  |
|  | [2, 3] | 1 | 2 | 1 |
|  | ≥4 | 2 |  |  |
| Limitations in running/jogging 1 kilometer | No | 0 | Yes | 3 |
|  | Yes | 3 |  |  |
| Limitations in walking 1 kilometer | No | 0 | No | 0 |
|  | Yes | 5 |  |  |
| Limitations in climbing several flights of stairs | No | 0 | Yes | 1 |
|  | Yes | 1 |  |  |
|  | Total points | 0-20 | Total points | 7 |
|  |  |  | Estimate of risk | 0.063 |

Notes: CHARLS, China Health and Retirement Longitudinal Study.

**Figure 4.**
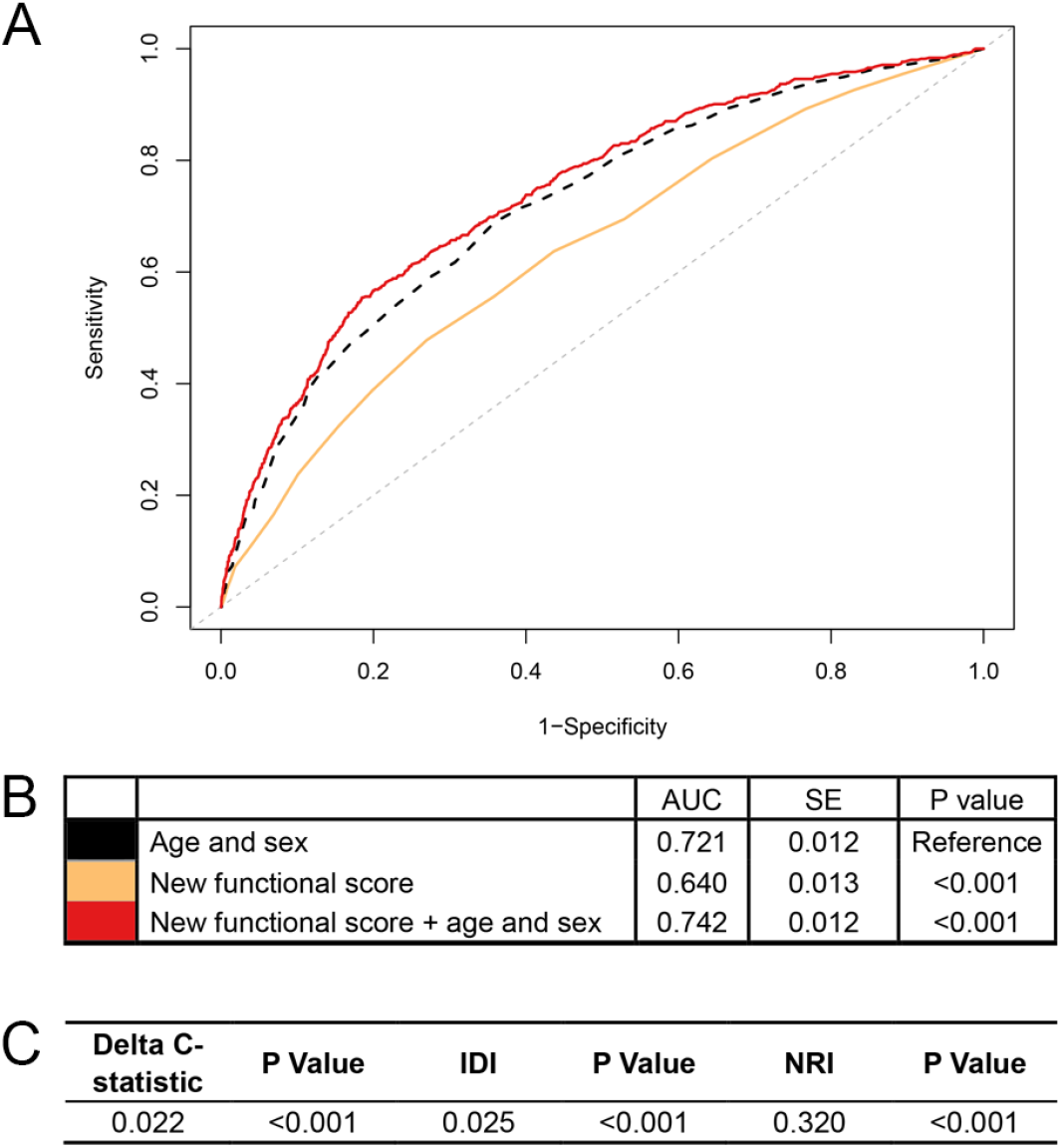
Association of the new functional score with all-cause mortality in CHARLS. Notes: CHARLS, China Health and Retirement Longitudinal Study; CI-PF, cognitive impairment and physical frailty; FI, frailty index; MCR, Motoric Cognitive Risk syndrome; AUC, area under the curve; SE, standard error; IDI, integrated discrimination improvement; NRI, net reclassification index. We calculated the continuous NRI and IDI using R package “PredictABEL”, in comparison to that of the basic model with age and sex. NRI equals to x% means that compared with persons without outcome, persons with outcome were almost x% more likely to move up a category than down. IDI equals to x% means that the difference in average predicted risks between the persons with and without the outcome increased by x% in the updated model. A shows receiver-operator characteristics curves for prediction of all-cause mortality for the new functional score. B shows the AUC for each model. C shows delta C-statistic, IDI, and NRI, in comparison to that of the basic model with age and sex.

In RAS, an independent dataset, we found that the new functional score predicted all-cause mortality risk as well, with an AUC of 0.618 (SE=0.026) (**Figure 5B**). More importantly, we found that the new functional score added predictive utility to the basic model with age and sex only. The AUC for mortality prediction was higher for a model with the new functional score, age, and sex (i.e., 0.689), relative to that of the basic model (i.e., 0.649). Adding the new functional score contributed significant improvements for predicting all-cause mortality in terms of reclassification, evidenced by the significant increase in IDI and continuous NRI relative to that of the basic model (all P<0.05, **Figure 5C**).

**Figure 5.**
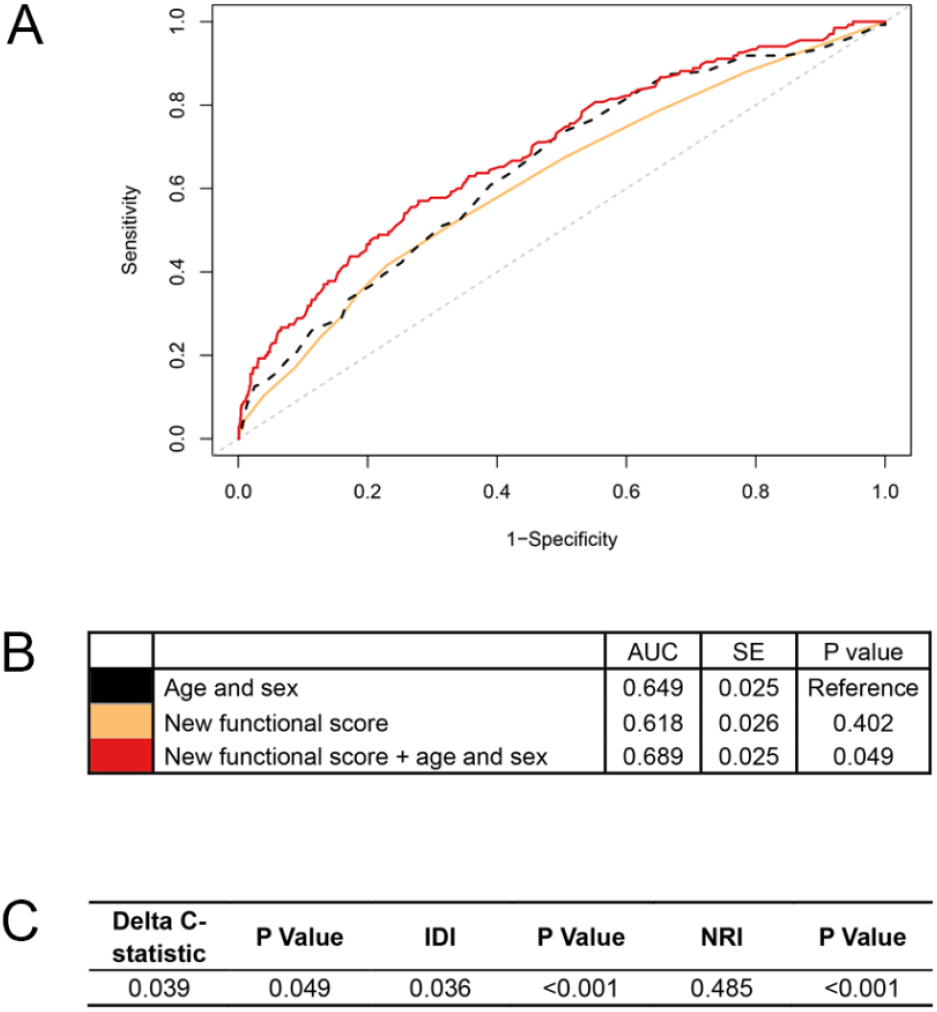
Association of the new functional score with all-cause mortality in RAS. Notes: RAS, Rugao Ageing Study; AUC, area under the curve; SE, standard error; IDI, integrated discrimination improvement; NRI, net reclassification index. We calculated the continuous NRI and IDI using R package “PredictABEL”, in comparison to that of the basic model with age and sex. NRI equals to x% means that compared with persons without outcome, persons with outcome were almost x% more likely to move up a category than down. IDI equals to x% means that the difference in average predicted risks between the persons with and without the outcome increased by x% in the updated model. A shows receiver-operator characteristics curves for prediction of all-cause mortality for the new functional score. B shows the AUC for each model. C shows delta C-statistic, IDI, and NRI, in comparison to that of the basic model with age and sex.

To help the public use of mortality prediction using the newly developed simple functional score, we provided an illustrative online tool (https://zipoa.shinyapps.io/mortalityprediction) based on parameters from CHARLS. In addition to the six self-reported items that we included in the new functional score, we also included age, sex, and education. We included age and sex as they are extremely important for health and are generally known to each person. We included education because of the same reason, and more importantly, because it may have some effects on the cognition-related item in the functional score (i.e., serial subtraction of 7 from 100). The integration then allows the user to get to know about his/her 6-year mortality risk after answering all items. For example, suppose a 60-year-old Chinese male, who graduated from middle school, could only count backward to 93 from 100 when doing the serial subtraction, had a BMI of 25 kg/m^2^, had hypertension and diabetes now, was limited in running 1 km and climbing several flights but was perfect in walking. Then he could get his 6-year mortality prediction of 13.1% from our simple online tool (**Figure 6**).

**Figure 6.**
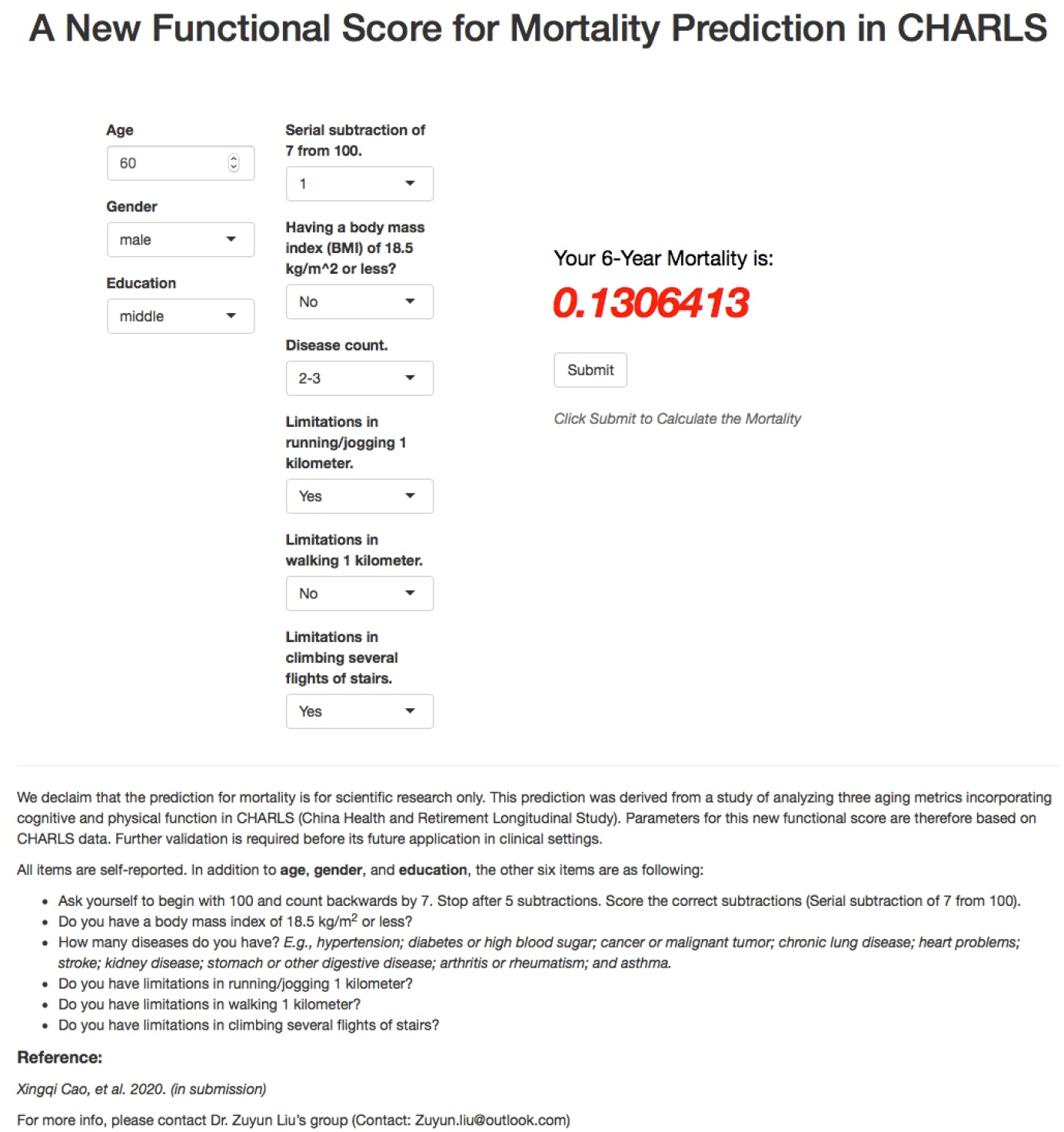
Illustration of mortality prediction for new functional score using the online tool for a 60-year-old Chinese person. Notes: CHARLS, China Health and Retirement Longitudinal Study.

## Discussion

Aging metrics incorporating cognitive and physical function are an emerging topic in gerontological and geriatric research, particularly due to its high predictive ability of poor prognosis. Various metrics have been developed, such as the three metrics mentioned in this study: CI-PF, FI, and MCR. There is a lack of understanding of how these metrics work when applying them in the same population in terms of identifying vulnerable persons and capturing the risk of adverse health outcomes. We showed that there was variability in the three metrics concerning the proportion of persons defined as vulnerable, and we provided a full picture showing how the three metrics capture mortality risk in two large national cohorts. Overall, these metrics were associated with mortality, with CI-PF and FI seeming to outperform MCR in mortality predictions. The findings indicated that various aging metrics incorporating cognitive and physical function could facilitate the early identification of vulnerable persons at risk, eventually reducing the burden of health care in aging societies with appropriate management.

We found that the three metrics incorporating cognitive and physical function are consistently associated with mortality risk in both Chinese and US persons. The stable results indicated that incorporation of cognitive and physical function can detect vulnerable persons at high risk of mortality, regardless of the various components of the three metrics and study populations. Previous studies have shown that integrating cognitive and physical function improves the prediction of adverse health outcomes, relative to the predictive utility of physical function alone [4], suggesting that cognition is crucial in geriatric functional assessments. Given that cognitive and physical functional decline could potentially be delayed [33, 34], and the fact that the aging metrics predict mortality as shown in our study, we therefore call for preventive and intervention programs aimed at ameliorating deteriorations in cognitive and physical function.

Despite the consistency of the main results, we noted the differences in the proportions of vulnerable persons identified by the three metrics. In the current study, the proportions of vulnerable persons defined by the CI-PF in CHARLS and NHANES were within that from previous studies, ranging from 1.0% to 12.1% among the community-dwelling persons [3, 11, 35]. However, the proportion of vulnerable persons defined by the FI was relatively higher than that of the CI-PF measurement. The great variability in the proportions of vulnerable persons is not surprising and may be due to many factors [36-39]. These factors include the age ranges and sex proportions of the sample, and the different setting of studies. For example, the discrepancy of the proportions of vulnerable persons defined by the MCR in CHARLS (19.6%) and NHANES (4.3%) could be attributed to the different age ranges in the two cohort studies.

Furthermore, the variabilities mentioned above imply that the three metrics may capture varying dimensions of functional aging. For instance, in CHARLS persons who were non-frail and pre-frail defined by FI, 26.2% and 36.2% were classified as cognitive impairment & non-frailty group for CI-PF, 10.6% and 23.3% were classified as MCR. This is expected because cognitive function is one of the many domains included in the FI measurement. The cognitive function of persons in the non-frail and pre-frail groups defined by FI score may be well maintained. The heterogeneity of our findings confirmed that different metrics combining cognitive and physical function cannot be assumed to be interchangeable.

Our results showed that CI-PF and FI have stronger mortality predictive utility than MCR in both Chinese and US persons in despite of the relatively low AUCs (partially due to the exclusion of some ill persons, e.g., those with ADL disability). This has not been reported previously. For instance, the AUC of CI-PF and FI for mortality prediction in CHARLS were 0.563 and 0.573, respectively, which were relatively higher than that of the MCR measurement (i.e., 0.507). Recent meta-analyses revealed that vulnerable persons defined by CI-PF or FI in older persons have a higher risk of all-cause mortality [40, 41]. Our study extends the findings of these previous studies. The PF, which is included in the CI-PF, is an objective metric (e.g., measurements of grip strength and gait speed), and is widely used in research because of its ability to predict adverse health outcomes (e.g., death [42, 43]). The better performance of the FI in predicting mortality risk could be attributed to the multidimensional and accumulative nature of the deficit approach, which could capture differences in the health status of persons at the same age [18]. These health deficits reflect multiple domains including comorbidities, cognition, psychology, symptoms, and disabilities [18]. The MCR is not a strong predictor of mortality relative to the other two metrics, intuitively because of its requirement to exclude persons with ADL disability. Theoretically, the MCR was designed to capture early signals of cognitive decline or functional changes occurring many years before the end of life.

However, when it comes to the clinical utility, CI-PF and FI may be inadvisable metrics. The CI-PF seems to have fewer items but has also been criticized for being impractical in clinical settings [44], because the CI-PF requires objective measures of cognitive impairment and PF, demanding much time and efforts. The multidimensional nature of FI measurement impedes its feasibility in large population studies. In addition to including sufficient numbers of deficits (at least 30), indeed, various FI scores differ significantly concerning their complexity as well as the stability in terms of adverse outcomes prediction [45]. Thus, we suggest that the feasibility and performance of the three metrics should be carefully balanced when using them.

As an example of illustrating how to balance the feasibility and performance, we developed a new simple functional score using six self-reported items from CI-PF and FI in CHARLS. The functional score comprised both cognitive (i.e., serial subtraction) and physical function domains (e.g., walking 1 kilometer). The mortality prediction by the functional score suggested that it could assist in identifying vulnerable persons at risk. Because of its self-reported nature and predictive utility for mortality risk, the functional score may be considered a simple and practical metric to assess functional deterioration.

This study has important implications. First, despite the heterogeneity of the three metrics, consistent associations with mortality were observed, supporting that cognitive and physical function have something in common, such as pathological mechanisms [46, 47]. This provides us with the opportunity to better track the future health trajectories of frail persons with cognitive impairment. Second, as introduced above, the new simple functional score does not require detailed physical examinations but relies on only six self-reported items, allowing it to be implemented in multiple settings. Finally, the predictive utility of these aging metrics including CI-PF, FI, and the new functional score supports the implementation of targeted interventions and health education at an early stage, which could effectively reduce mortality risk at a lower cost. With appropriate management, it is expected to alleviate the burden of health care in those with varying cognitive and physical status.

Our study has several strengths. First, the two cohorts included in our study are representative samples from two of the largest countries in the world, which substantially differ in many aspects such as social-economic position and lifestyle. The consistent results from the two cohort studies strengthened our findings. Second, we presented the three aging metrics in the same population and examine their associations with mortality, which is scarce in the literature. Nevertheless, several limitations of this study should be mentioned. First, the CHARLS has a relatively short follow-up period (i.e., up to 6 years), impeding us to evaluate the long-term effect of cognitive and physical impairment on adverse health outcomes in this cohort. Second, it should be noted that there are differences between study samples of CHARLS and NHANES, such as ethnic differences and age ranges. These may lead to variation in results between the two samples (e.g., proportions of vulnerable persons identified by the three metrics). Third, there were differences in the components of the three metrics across the two cohorts. Nevertheless, the metrics have been proved to be valid in previous studies [11, 28, 48, 49]. Also, the three metrics are different in terms of target population and initial purpose. Thus, applying them in different settings should be done with caution. Finally, the new functional score was validated only in the Chinese population. More studies are required to repeat our analyses in the US and other countries.

In both Chinese and US persons, we found that aging metrics incorporating cognitive and physical function consistently capture mortality risk, despite their inherent substantial differences. The incorporation of cognitive and physical function has the potential for risk stratification in both research and clinical settings. The findings support the implementation of preventive strategies and intervention programs targeting at these metrics to improve the quality of life and further reduce premature death. The new functional score we developed can predict mortality risk as well, showing good feasibility and performance. Nevertheless, it requires more studies involving different population samples and examining associations of this new score with other age-related outcomes.

## Data Availability

The CHARLS data are publicly available through the CHARLS website at http://charls.pku.edu.cn/en. The NHANES data are available for sharing within the scientific community. Researchers can apply for data access at https://www.cdc.gov/nchs/nhanes/index.htm. The RAS data are available on request from the corresponding authors (XW and ZL).

https://www.cdc.gov/nchs/nhanes/index.htm

## Author contributions

ZL conceived and designed the study. XC and CC performed the analysis and wrote the initial draft of the manuscript. ZJ, QLX, EOH, XL, SL, XW, YZ, and ZL helped to interpret the results and edit the manuscript. QLX, EOH, XW, YZ, and ZL contributed to the critical revision of the manuscript for important intellectual contents. All authors reviewed and approved the final version of the manuscript.

## Acknowledgments

We thank all respondents of the China Health and Retirement Longitudinal Study (CHARLS), the US National Health and Nutrition Examination Survey (NHANES), and the Rugao Ageing Study (RAS) for their participation.

